# Detection of autoimmune antibodies in severe but not in moderate or asymptomatic COVID-19 patients

**DOI:** 10.1101/2021.03.02.21252438

**Authors:** Aisha D. Fakhroo, Gheyath K. Nasarallah, Taushif Khan, Farhan S. Cyprian, Fatima Al Ali, Manar M.A. Ata, Sara Taleb, Ali A. Hssain, Ali H. Eid, Laith J. Abu-Raddad, Abdullatif Al-Khal, Asmaa A. Al Thani, Nico Marr, Hadi M. Yassine

## Abstract

The heterogeneity of COVID-19 lies within its diverse symptoms and severity, ranging from mild to lethal. Acute respiratory distress syndrome (ARDS) has been shown to be the leading cause of mortality in COVID-19 patients, characterized by a hyper cytokine storm. Autoimmunity is proposed to occur as a result of COVID-19, given the high similarity of the immune responses observed in COVID-19 and autoimmune diseases. Here, we investigate the level of autoimmune antibodies in COVID-19 patients with different severities. Initial screening for antinuclear antibodies (ANA) IgG revealed that 1.6% (2/126) and 4% (5/126) of ICU COVID-19 cases developed strong and moderate ANA levels, respectively. However, all the non-ICU cases (n=273) were ANA negative. The high ANA level was confirmed by immunofluorescence (IFA) and large-scale autoantibody screening by phage immunoprecipitation-sequencing (PhIP-Seq). Indeed, the majority of the samples showed “speckled” ANA pattern by microscopy, and we demonstrate that samples of ICU patients with strong and moderate ANA levels contain autoantibody specificities that predominantly targeted proteins involved in intracellular signal transduction, metabolism, apoptotic processes, and cell death; further denoting reactivity to nuclear and cytoplasmic antigens. In conclusion, our results further support the notion of routine screening for autoimmune responses in COVID-19 patients, which might help improve disease prognosis and patient management. Further, results provide compelling evidence that ANA-positive individuals should be excluded from being donors for convalescent plasma therapy in the context of Covid-19.

## 1 Introduction

The severity of COVID-19 is diverse, with a wide range of symptoms, characterized by mild to lethal. Upon viral infections, a robust immune response is usually initiated involving both the innate and adaptive immune system in order to irradicate the virus [1]. Typically, the viruses evade these immune responses through one of the following mechanisms; (1) Molecular Mimicry, (2) bystander activation, and (3) epitope spreading [1]. In COVID-19 patients, several immunological impacts have been observed including, hyper-immune response, abnormal cytokine/chemokine production, T cells hyperactivation, increased monocytes, macrophages and neutrophils count [1]. In fact, the cytokine storm (i.e., abnormal cytokine secretion) has been shown to be the leading cause of death in COVID-19 patients, where they usually experience a hyper pro-inflammatory immune response leading to ARDS [1]. These COVID-19’s immune responses are quite similar to those observed in autoinflammatory and autoimmune conditions [1]. It is known that infectious diseases trigger autoimmunity, specifically through molecular mimicry, and several viruses have been associated with autoimmune diseases [1,2,3]. For example, enteric viruses have been associated with type 1 diabetes, and herpesviruses infections have led to the development of several autoimmune disorders (e.g., multiple sclerosis, systemic lupus erythematosus, and rheumatoid arthritis) [1,3]. In addition, mice infected with murine coronavirus developed immune-mediated encephalomyelitis [4]. In another study, rhinovirus and coronavirus were shown to be the highest frequently detected pathogens in patients with psoriasis flares following respiratory tract infections [5].

In terms of COVID-19, severe cases have been suspected of causing autoimmunity. Several reports have suggested a link between SARS-CoV-2 infection and Kawasaki disease, an acute inflammation of the blood vessels affecting children [2,6]. Moreover, patients with critical COVID-19 pneumonia reported neutralizing IgG autoantibodies against type I IFNs [7]. Similarly, here we suggest that critically ill COVID-19 patients may experience an elevated level of antinuclear antibodies (ANA) [8]. High levels of ANA have been previously associated with many autoimmune disorders, such as SLE and lupus [8]; however, it has not been linked to COVID-19 yet. Therefore, this study investigates whether COVID-19 leads to autoimmunity through measuring ANA level in blood sera samples collected from COVID-19 patients with different clinical severities (i.e., ICU “Severe” vs. Non-ICU “Mild or Asymptomatic”). Here, we report a high level of ANA in some severe COVID-19 cases, suggesting a possible contribution of COVID-19 to autoimmunity, and therefore, exacerbated disease outcome.

## 2 Method

### 2.1 Sample collection and ethical compliance

This study was approved by the IRB committees of Hamad Medical Corporation (MRC-01-20-145) and Qatar University (QU-IRB 1289-EA/20). Sera samples were collected from COVID-19 patients at different clinical stages classified into two groups; (1) mild/asymptomatic (Non-ICU, n=273), (2) Severe/Critical (ICU, n=126). Out of the 126 ICU patients, only 80 patients had sera extracted at different time points with the assumption that ICU admission is Day 1.

### 2.2 ANA IgG ELISA assay

Sera samples were diluted 1:101 in sample diluent (DILSPE), and ELISA was performed using ANA Screening IgG kit (DIA.PRO, Italy) according to the manufacturer standards. The cutoff value was calculated as follows: Negative Control (OD450nm) + 0.250. Samples with cut of value (S/Co) <0.8, S/Co (0.8-1.1), S/Co>1.1 were considered negative “normal”, equivocal “moderate” and positive “abnormal” respectively. Note that samples tested positive and equivocal were repeated in triplicates with mean and standard deviation calculated.

### 2.3 ANA HEp-2 IFA assay

Indirect Fluorescent Antibody (IFA) technique was performed using AccuDiag™ ANA HEp-2 IFA kit (Diagnostic Automation/Cortez Diagnostics, US) according to the manufacturer standards. A positive reaction is indicated by the presence of any pattern of nuclear apple-green staining observed at a 1:40 dilution based on a 1+ to 4+ scale of staining intensity (+1 weak → +4 strong).

### 2.4 Phage Immunoprecipitation-Sequencing (PhIP-Seq) and peptide enrichment analysis

PhIP-Seq and peptide enrichment analysis were performed as previously described [9] using the T7 Human ORF 90mer library, a phage display library expressing 90-aa protein fragments tiling through the human proteome with a 45-aa overlap [10]. The T7 Human ORF 90mer Library had been obtained from S. Elledge (Brigham and Women’s Hospital and Harvard University Medical School, Boston, MA, USA). In brief, we imputed −log_10_(*P*) values as described previously [9] by fitting a zero-inflated generalized Poisson model to the distribution of reading counts obtained from the tested samples following immunoprecipitation and regressed the parameters for each peptide sequence based on the read counts obtained from an input library sample (*i*.*e*., prior to immunoprecipitation). These −log_10_(*P*) values were considered as peptide enrichment scores and reflected a quantitative measure for the presence of an autoantibody specificity in a given sample. A peptide enrichment score of ≥ 2.3 was considered to be statistically significant. We also removed peptides from the downstream analysis that were enriched in mock-IP samples, which served as negative controls, and only considered peptides that were significantly enriched in at least two test samples. We then computed log odds ratios (lod) for all retained peptides (n = 328) in order to identify autoantibody specificities that were differentially enriched between ICU patients and asymptomatic COVID-19 cases. Peptides with a |lod| ≥ ln(1.5) were considered differentially enriched (n = 79).

### 2.5 Gene set enrichment analysis

Out of the 79 differentially enriched peptides, 62 were derived from coding sequences with a defined gene annotation (Entrez) and were considered for gene enrichment analysis. For this analysis, we used the Molecular Signatures Database (MSigDB) as previously described [11]. Out of the 62 queried genes, 42 were found to be significantly enriched (*P*-value < 10^−5^ and FDR q-value < 0.05) in one of 20 gene sets (**Figure 3C**).

**Figure 1.**
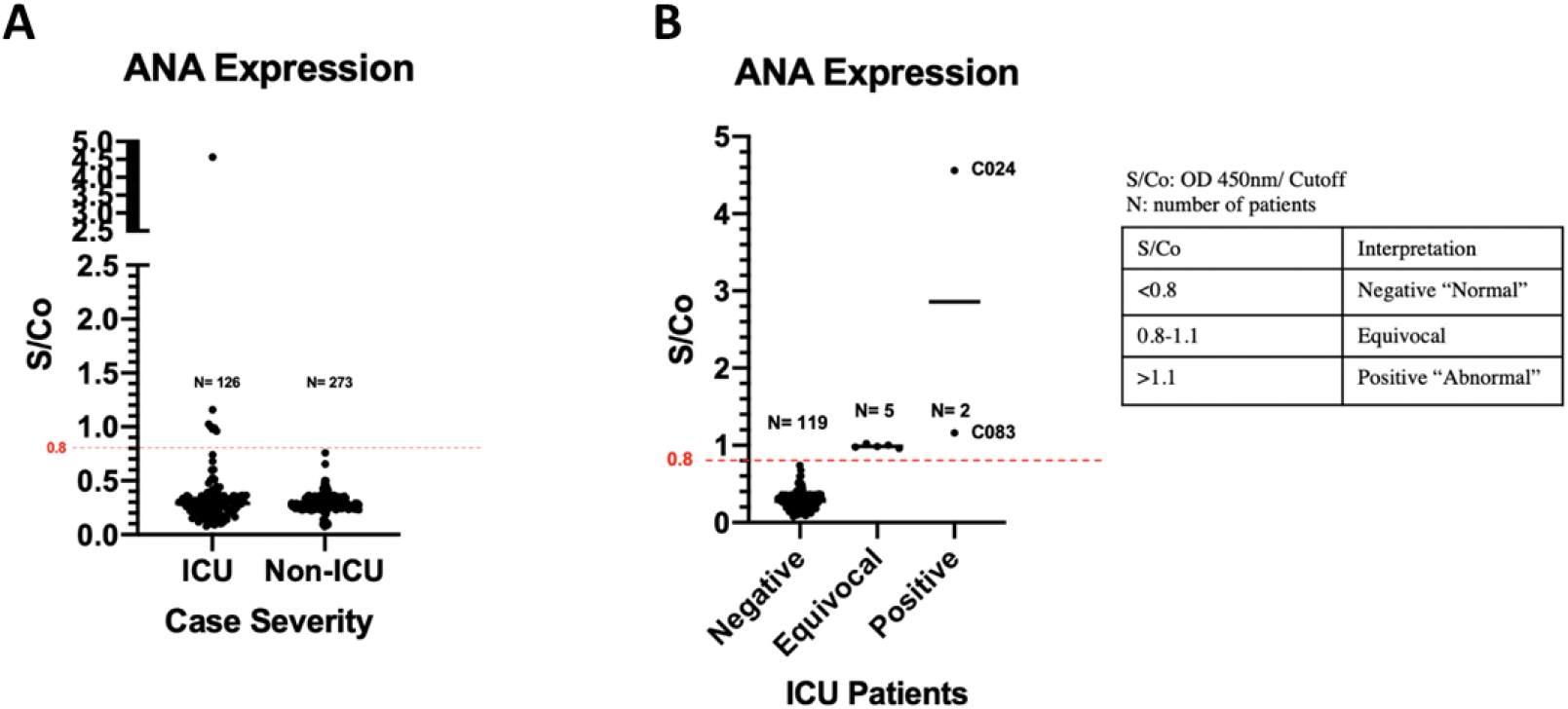
ANA ELISA levels of the COVID-19 Patients. **(a)** sera samples of ICU (n=126) and non-ICU (n=273) COVID-19 patients; and **(b)** sera samples of ICU COVID-19 patients (n=126). Samples were tested at 1:101 dilution.

**Figure 2.**
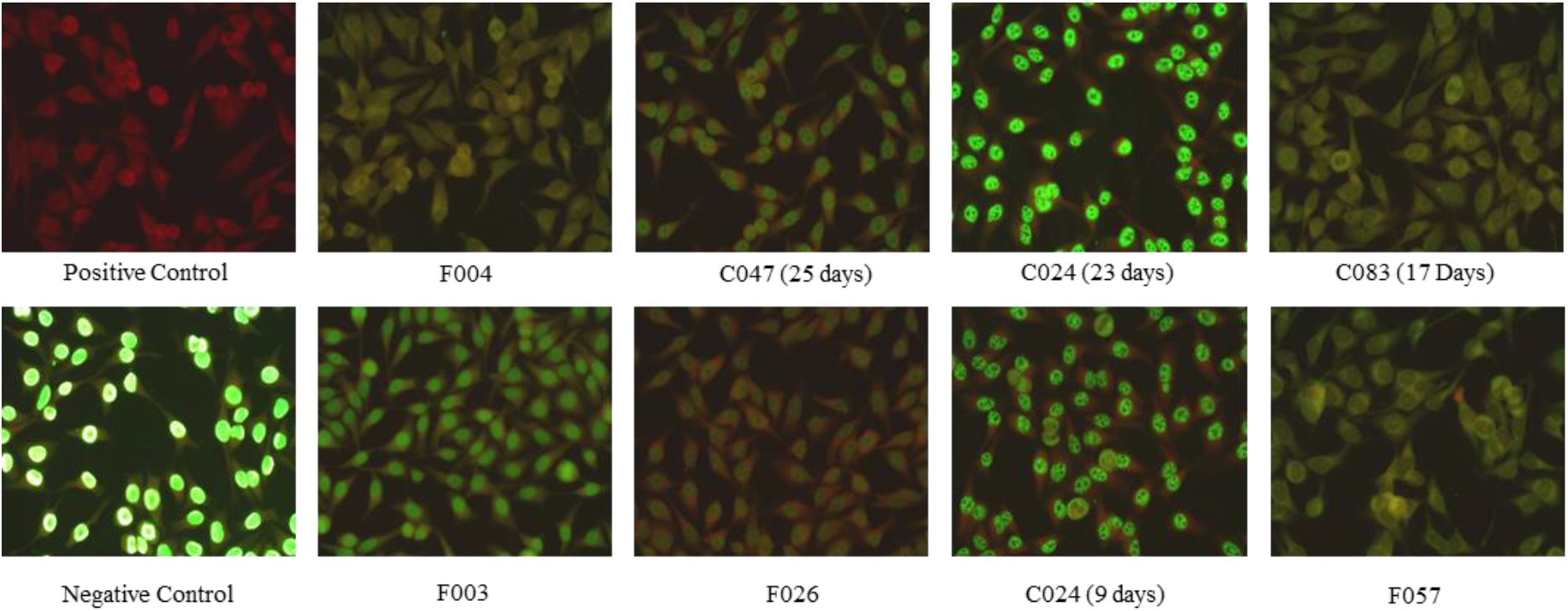
Immunostaining of Hep2 Cells with ANA from COVID-19 Sera Samples. Confirmation of ANA level in positive and equivocal samples (F003, F004, F026, C047 “25 days”, C024 “9 days”, C024 “23 days”, F057 and C083 “17 days”) using IFA technique, where HEp2 cells were used as a substrate to detect ANA antibodies in human serum. Samples were tested at 1:40 dilution. Note that # of days corresponds to the time of sample collection assuming ICU admission is day 1.

**Figure 3.**
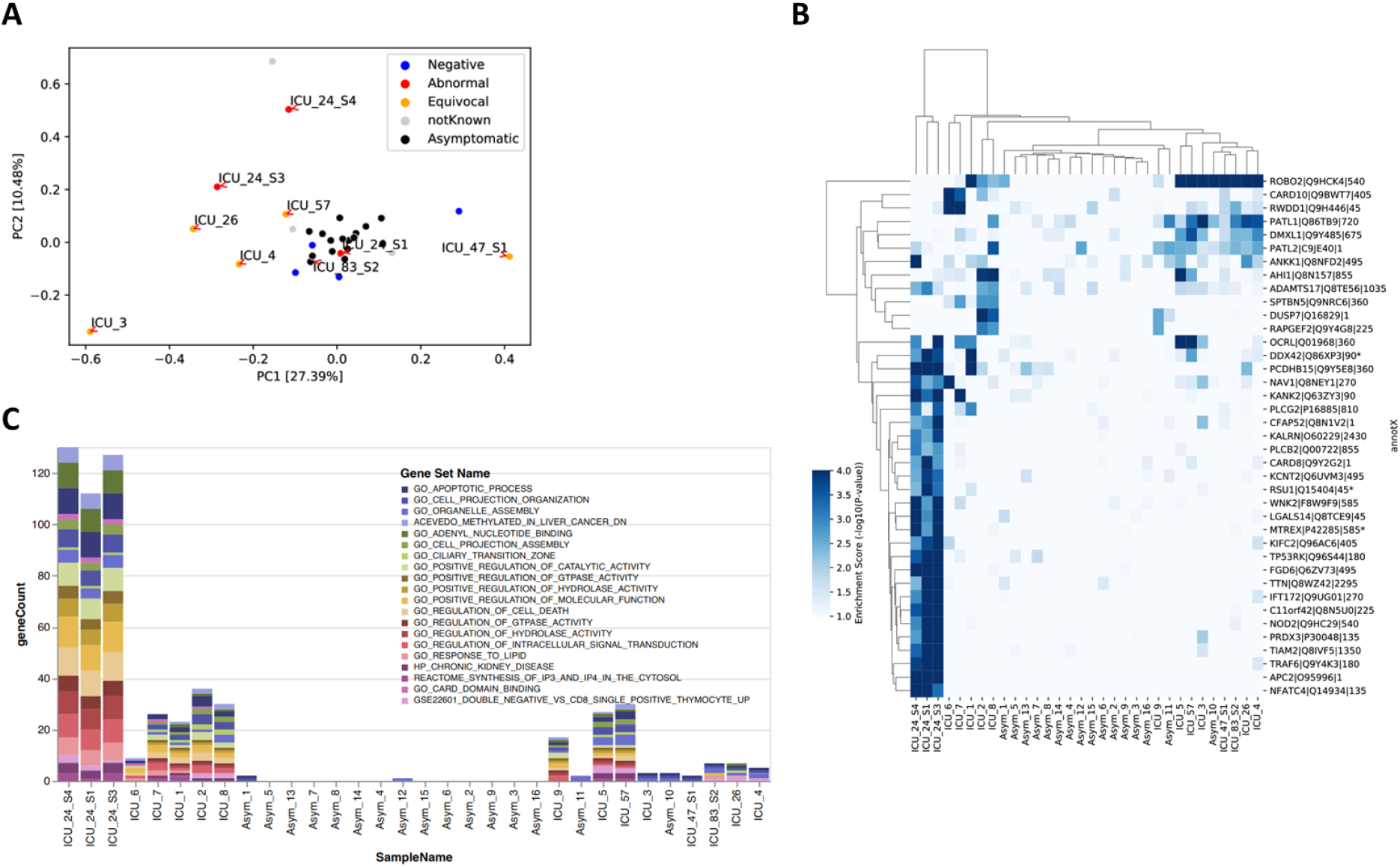
Autoantibody profile of selected cases assessed by PhIP-Seq. **A**, Principal component analysis of the peptide enrichment scores reflecting autoantibody-autoantigen interactions in ICU patients and asympromatic COVID-19 cases. The color core indicates the ANA status. **B**, Heatmap plot showing the binding profile of the 79 differentially enriched (DE) peptides in ICU cases versus asymptomatic cases, with hierarchical clustering. Each row indicates a human peptide (90mer, start position is indicated relative to the UniProtKB entry), each column represents a sample. The color gradient for each cell of the heatmap plot represents the peptide enrichment score (−log_10_(*P*) value) for a given antigenic peptide and sample. A -log_10_(*P*) ≥ 2.3 was considered significantly enriched; * Represtns known autoantigens with an entry in the human autoantigen database (AAgAtlas 1.0). **C**, Stacked bar plot showing the results of a gene set enrichment analysis of the peptides shown in (**B**). The color code indicate the gene sets from the the Molecular Signatures Database (MSigDB) for which at least one DE peptide was enriched (*P*-value < 10^−5^ and FDR q-value < 0.05). Samples are sorted as shown in (**B**) according to hierarchical clustering.

### 2.6 Detection of shared linear B cell epitopes

To test for shared linear B cell epitopes between the identified autoantigens and SARS-CoV-2 antigens, we built a pairwise distance matrix that captured the maximum size of linear sequence identity of amino acids between the 79 differentially enriched human 90 aa peptides and 17 reference sequence of SARS-CoV2 proteins in UniProtKB (https://covid-19.uniprot.org) as previously described [12]. A linear sequence identity of 7 amino acids or more was considered a shared linear B cell epitope.

## 3 Results

ANA antibodies were screened in non-ICU (n=273) and ICU (n=126) COVID-19 patients using ANA IgG ELISA. All of non-ICU patients (n=273) tested negative; (S/Co<0.8) with an average of 0.286 (+/-0.073). On the other hand, 7/126 ICU patients (∼5.6%) reported a S/Co value of >0.8, showing moderate-high ANA level (**Figure 1a**). 4% of ICU patients (5/126) tested equivocal (S/Co: 0.8-1.1, moderate ANA level), whereas 1.6% (2/126) tested positive (S/Co>1.1, abnormal ANA level) (**Figure 1b**). The positive samples, C024 and C083 had a S/Co value of 4.561 and 1.159, respectively (**Figure 1b**). To confirm the presence of ANA, IFA was performed where HEp-2 cells were immunostained with ANA from sera samples (Positive and Equivocal) (**Figure 2**). The majority of the samples (C024, F026, F004, C047) showed a “speckled” ANA pattern (**Figure 2**). In addition to ANA level, cytoplasmic ANA level was observed in samples F003, F004, F057, and C083 (**Figure 2**). Samples F057 and C083 showed a “punctate nuclear envelope” ANA pattern (**Figure 2**). Note that all of the patients tested equivocal/positive for ANA are males within the age range of 41-75 years (Average= 55 years) (**Table 1**). In relation, 71.4% of patients (5/7) are co-diagnosed with hypertension, and the mortality rate was 42.9% (3/7), specifically samples F003, F026, and F057 that showed moderate ANA Level (**Table 1**). Diabetes was also common, accounting for 42.8% of patients (3/7), including patient C083, which exhibited abnormal ANA levels. Finally, we performed a large-scale autoantibody screen of the ANA-positive (C024, C083) and five equivocal sera samples using phage-immunoprecipitation sequencing ^[9]^. Selected ANA-negative samples obtained from ICU patients with COVID-19 (n = 7) and from asymptomatic COVID-19 cases (n = 15) were assayed for comparison (**Supplementary Table S1**). Principal component analysis of the peptide enrichment scores confirmed that most ANA-positive and equivocal samples of ICU cases clustered separately from the asymptomatic COVID-19 cases, with the exception of the day 9 sample of C024 and the sample collected from C83 on day 17 after ICU admission (**Figure 3A**). We identified 79 autoantibody specificities that were differentially enriched in ICU patients in comparison to the asymptomatic COVID-19 cases (**Supplementary Table S1**), allowing a clear separation of critical versus asymptomatic COVID-19 cases by hierarchical clustering (with the exception of one asymptomatic case) (**Figure 3B**). Interestingly, our unbiased screen revealed autoantibody specificities against several known autoantigens present in AAgAtlas 1.0, a human autoantigen database [13], including nuclear proteins such as the DEAD-Box Helicase 42 (DDX42) and Mtr4 Exosome RNA Helicase (MTREX), as well as proteins involved in immune defences and cellular signaling, such as Immunoglobulin Superfamily DCC Subclass Member 1 (DCC) and Ras Suppressor Protein 1 (RSU1). To functionally characterize the self-antigens that were being targeted in critically ill COVID-19 patients, we performed a gene set enrichment analysis using the Molecular Signatures Database (MSigDB) ^[11]^ and including all of the 79 putative autoantigens for which we had found autoantibodies to be differentially enriched among the tested ICU cases when compared to asymptomatic COVID-19 cases. This analysis confirmed that autoantibodies in ICU patients primary targeted intracellular proteins involved in intracellular signal transduction, metabolism, apoptotic processes, and cell death. Autoimmune responses were primarily observed in samples with moderate and high ANA levels, particularly in the samples of patient C024 with the highest ANA measurements, which appeared to increase over time (**Figure 3C**).

**Table 1:**
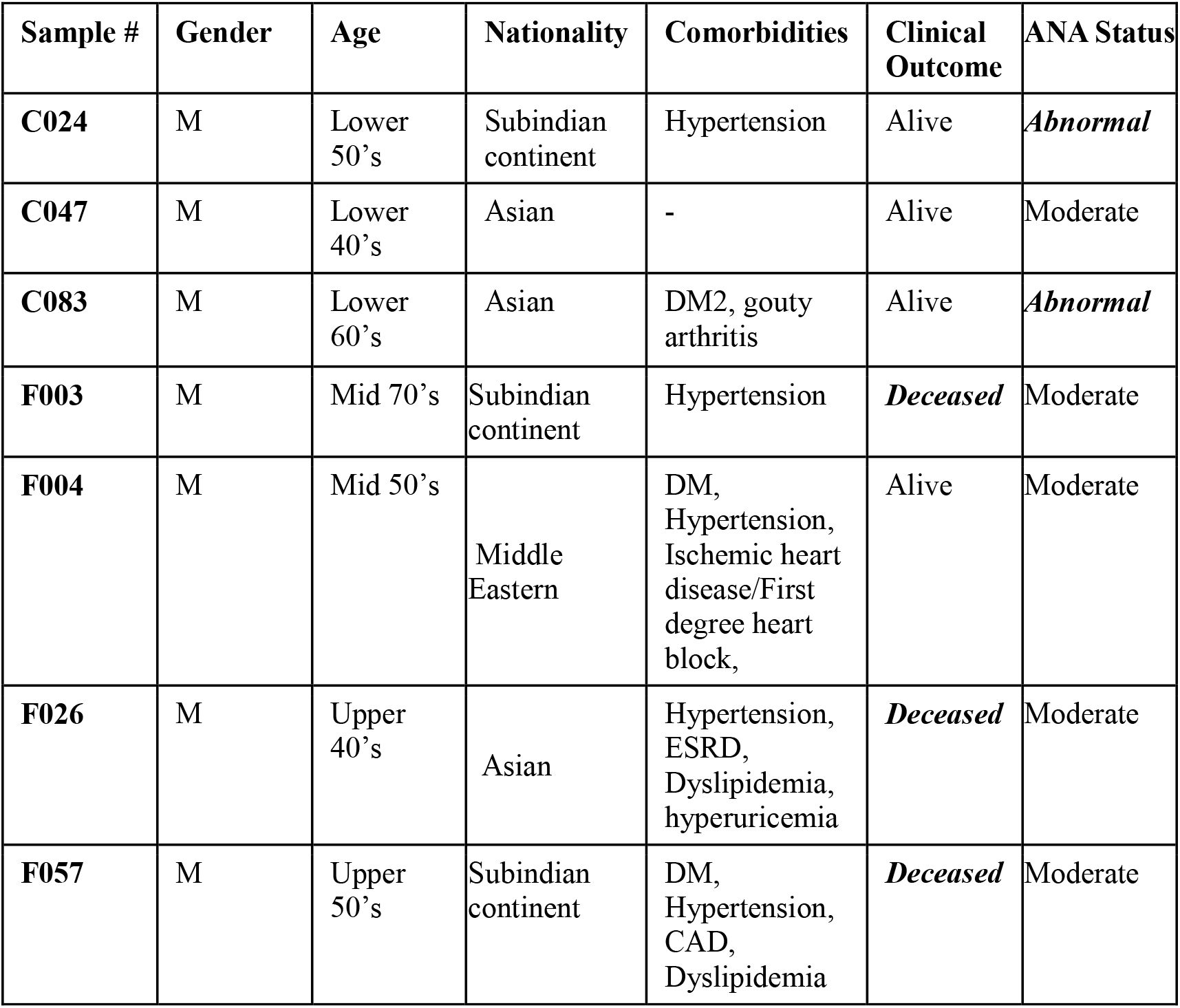
Demographic Data of COVID-19 Patients with Moderate-to-Abnormal ANA Level.

## 4 Discussion

COVID-19 disease progression may pass through up to four different phases [1]. The first phase is characterized by an initial viral infection phase that is usually mild or asymptomatic in approximately 80% of patients [1]. The host-virus interactions then delineate the progression of the disease. Some patients progress to a second phase, characterized by a hyper-immune response (i.e, cytokine storm) [1]. A state of hypercoagulability occurs in the third phase [1]. In combination, they may lead to organ damage in the fourth phase, which is usually mediated by the host’s innate immune system [1,6]. In this present study, we assessed the generation of antinuclear autoimmune antibodies (ANA) in critically ill COVID-19 patients in order to gain a better understanding of the disease prognosis and to pave the way for the possible use of immunomodulatory drugs for the treatment of these patients. It is worth noting that several immunomodulatory drugs have been proved to be effective in relieving COVID-19 symptoms, such as tocilizumab [1].

Interestingly, ANA were exclusively observed in ICU COVID-19 patients (7/126, 5.6%), which suggests a potential correlation between COVID-19’s severity and ANA production. In other words, SARS-CoV-2 infection may have triggered the production of ANA autoantibodies leading to possible cases of autoimmunity in severely ill COVID-19 patients. However, the mechanism is yet to be studied. In general, viral infections trigger autoimmunity through one of the following mechanisms, (1) molecular mimicry, (2) bystander activation, and (3) epitope spreading [3]. In molecular mimicry, viruses display antigens structurally similar to self-antigens activating a cross-reactive immune response against both self and non-self-antigens. During bystander activation, a non-specific hyper anti-viral immune response characterized by a pro-inflammatory environment causes the release of self-antigens from damaged tissues, which are then presented by antigen-presenting cells (APC), triggering autoreactive T cells and autoimmunity. One such example is HIV, which mimics human T-cell receptor (TCR) to a high extent where autoantibodies are produced [14]. In support of this mechanism, our large-scale autoantibody screen of selected patients by PhIP-Seq revealed several known and novel autoantigens among ICU cases, particularly in the ICU patient with moderate and high ANA responses. An in-depth analysis of these autoantibody specificities confirmed that these autoimmune responses were primarily directed against intracellular proteins and, therefore likely as a consequence of extensive tissue damage during disease progression. Of note, the patient with the highest ANA serum levels and most robust autoantibody responses as assessed by PhIP-Seq (a 53 year old male with Indian nationality) had a clinical history of hypertension, but was otherwise previously healthy. Similarly, epitope spreading is characterized by the release of more self-antigens activating autoreactive T cells that eventually spread to other autoreactive T cells. A recent study identified cross-reactive epitopes between SARS-CoV-2 and human molecular chaperones [15]. Bioinformatics analysis showed that a family of heat shock proteins (Hsp70) shared antigenic epitopes with SARS-CoV-2, capable of inducing autoimmunity against endothelial cells through the process of molecular mimicry [15]. Thus, a similar mechanism may apply to ANA production, where cross-reactive epitopes between SARS-CoV-2 and nuclear antigens may exist. However, none of the autoantigens we have identified in this present study shared linear B cell epitopes with any of the SARS-CoV-2 protein reference sequences (**Supplementary Figure S1**).

In terms of comorbidities, hypertension was common among patients that showed ANA level, specifically in those who were deceased (F003, F026, and F057). Thus, this may suggest a potential link between hypertension and ANA level. However, a more extensive cohort study is needed to validate this hypothesis. Hypertension is prevalent in several autoimmune diseases, including SLE and RA promoting chronic inflammation [16]. Here, we speculate that hypertension may act as a risk factor promoting a pro-inflammatory environment, possibly leading to autoimmunity through bystander activation. Although these samples showed moderate ANA levels, it is inconclusive because extra time points of the samples are needed to check ANA level change over time. The second most common comorbidity was diabetes, specifically patient C083, who experienced both type 2 diabetes and abnormal ANA level. Type 2 diabetes is suspected to be an autoimmune condition given the presence of circulating autoantibodies against β cells [17]; thus, this may contribute to abnormal ANA levels (i.e., risk factor). Furthermore, ANA level was confirmed using IFA, where HEp-2 cells were immunostained with ANA expressed in sera samples. ANA level comes in different patterns depending on the specific antigens to which ANA bind. According to the results, most of the positive/equivocal samples showed speckled patterns suggesting potential antigens such as n-RNP, Sm, and SSB/La [18].

In a previous ANA screening (i.e., pre-pandemic) of 2655 randomly selected samples from blood bank donors (unpublished results), ten samples showed abnormal ANA levels (∼0.38%). Among these positive ANA samples, 5 (50%) were positive for other viral infections (Table 2). Four samples (A, B, C, and D) tested positive for B19 Virus IgG, two samples (A and E) for WNV (West Nile Virus) IgG, and one sample (A) for Dengue virus. B19 virus has been associated with several autoimmune diseases, such as rheumatoid arthritis, systemic lupus, antiphospholipid syndrome, systemic sclerosis, and vasculitides [19]. It has also been shown to induce cross-reactive autoantibodies by means of molecular mimicry between parvovirus VP1 protein and host proteins (e.g., human cytokeratin and transcription factor GATA 1) [19]. Likewise, WNV infection has been reported to promote autoimmune conditions, including myasthenia gravis (MG), through the process of molecular mimicry [20]. In addition, several studies have shown cross-reactivity between antibodies directed against dengue virus nonstructural protein 1 (NS1) and human platelets/endothelial cells damaging them [21]. As observed, molecular mimicry and autoimmunity are common among these viruses, which may suggest a similar mechanism taking place during SARS-CoV-2 infection.

**Table 2:** Screening of Positive ANA Prepandemic Samples (i.e., prior COVID-19) Against Different Viruses.

| Sample | Gender | ANA IgG | Dengue | Chikungunia | HEV IgM | HEV IgG | B19V IgM | B19V IgG | WNV IgM | WNV IgG |
| --- | --- | --- | --- | --- | --- | --- | --- | --- | --- | --- |
| A | M | positive | positive | Negative | ND | ND | Negative | Positive | Negative | Positive |
| B | M | positive | Negative | Negative | Negative | Negative | Negative | Positive | Negative | Negative |
| C | M | positive | ND | ND | Negative | Negative | Negative | Positive | ND | ND |
| D | M | positive | ND | ND | ND | ND | Negative | Positive | ND | ND |
| E | M | positive | ND | ND | ND | ND | ND | ND | Negative | Positive |

ND: Not determined.

In conclusion, this study sheds light on a potential relationship between COVID-19 and autoimmunity, particularly ANA production. Nevertheless, some limitations are associated with this study. First, the sample size is relatively small, and a larger-scale study may be needed. Second, some of the sera samples were only taken at one time point, which makes it harder to explore ANA change over time. Third, people are admitted to ICU mostly after 7 days after infection, so the time points of the ICU samples are calculated from their admission to ICU rather than the beginning of the infection. Thus, the acute vs. convalescent phase inconsistency of the ICU and non-ICU samples may affect the accuracy of the results. Still, this study provides a closer insight into the immunological progression of the disease and its prognosis. We, therefore, propose to include the screening for autoimmune antibodies as a routine test for COVID-19 patients.

## Data Availability

Data will be available upon a direct request from the corresponding author.

## Authors contribution

HY and GN conceptualized the idea. FC, AH, AA and ST provided samples. AF performed ELISA and microscopy experiments and wrote the first draft of the paper. FA and MA performed the PhIP-Seq experiments. TK and NM analyzed data and revised the manuscript. HY, GN, NM and AAA provided funding. All authors revised the manuscript and approved it before submission.

## Funding

This project was supported with funding from Qatar National Research Funds (QNRF), grant #NPRP11S-1212-170092, and funding from Sidra Medicine (SDR400048).

## Conflict of interest

The authors have declared that no conflict of interest exists.

## Ethical approval

This study was approved by IRB committees of Hamad Medical Corporation (MRC-01-20-145), Sidra Medicine (IRB Protocol #: 1511001953) and Qatar University (QU-IRB 1289-EA/20)

## Acknowledgment

The authors would like to thank Ms. Reham Ghazal and Ms. Maria Smatti for their help in samples collection and processing, Stephen Elledge (Brigham and Women’s Hospital, Harvard University Medical School) for kindly providing the T7 Human PRF 90mer Library used in this study and the Integrated Genomics Services team of Sidra Genomics for their assisting processing the custom sequencing library pools.

Data will be shared upon a direct request to the corresponding author.

---

**Supplementary table 1.**
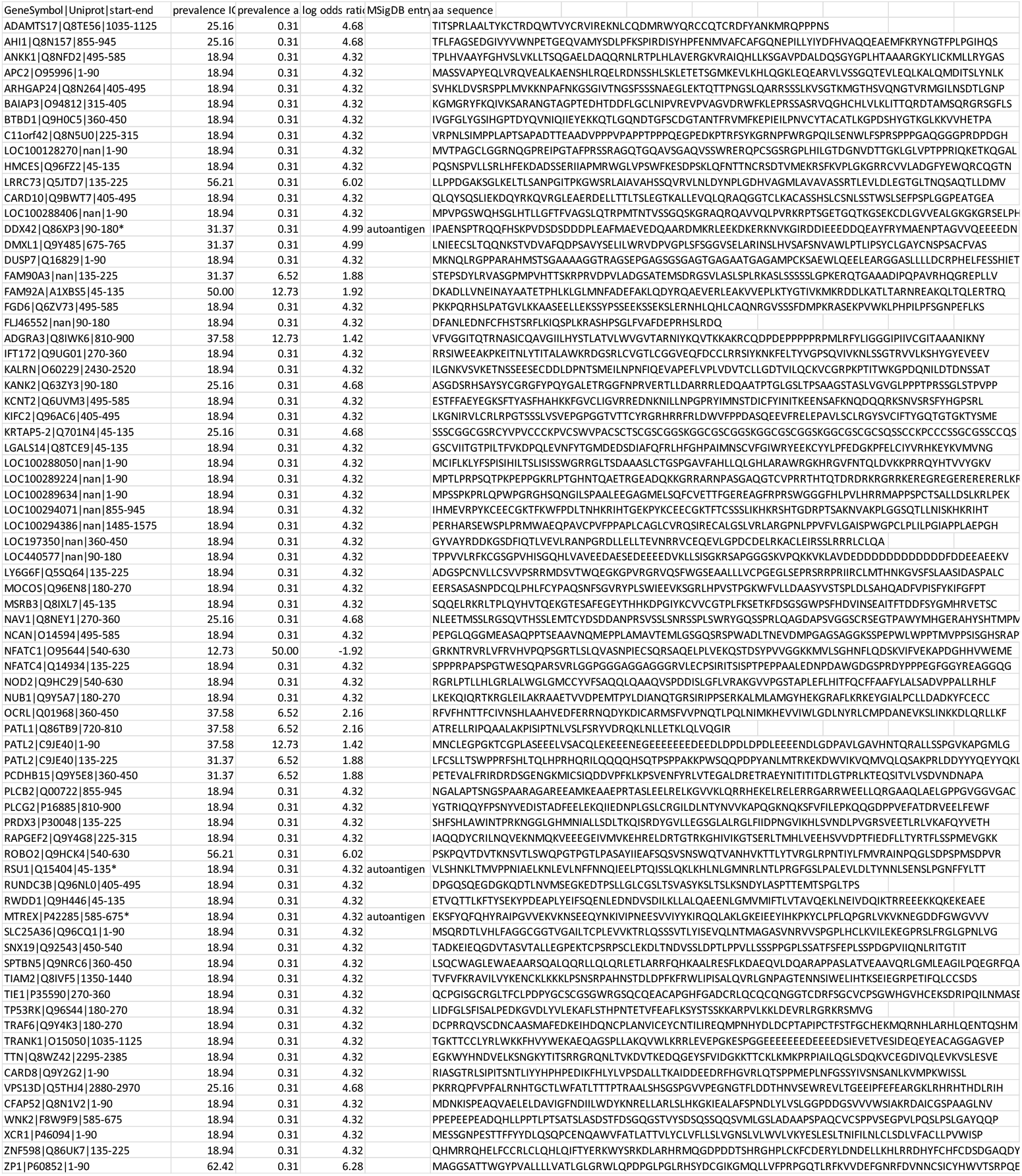
Identified autoantibody specificities that were differentially enriched in ICU patients.

**Supplementary figure 1:**
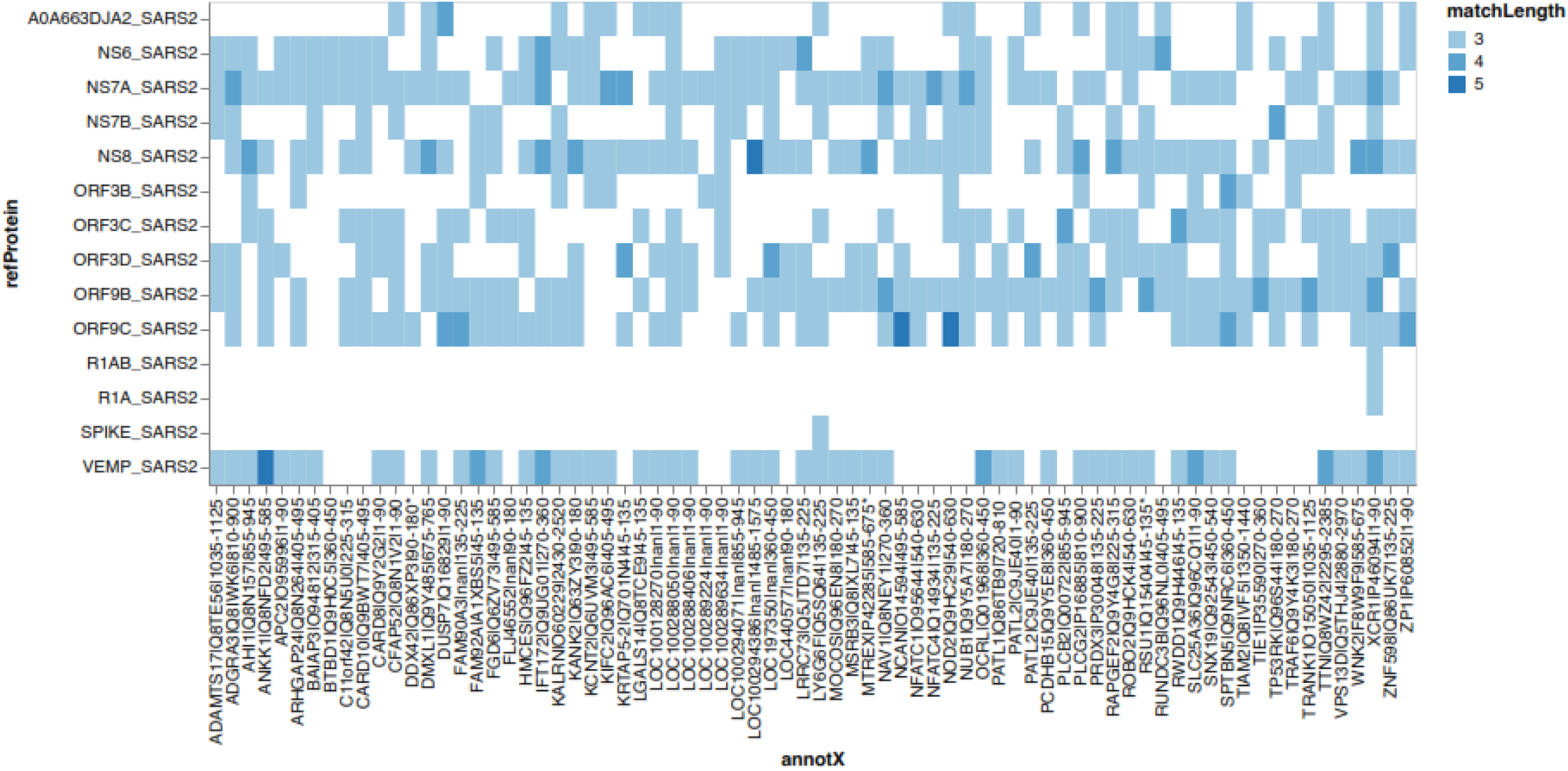

## Notes

### Competing Interest Statement

The authors have declared no competing interest.

### Author Declarations

Ethical approval This study was approved by IRB committees of Hamad Medical Corporation (MRC-01-20-145), Sidra Medicine (IRB Protocol #: 1511001953) and Qatar University (QU-IRB 1289-EA/20)

